# High vaccine effectiveness against COVID-19 infection and severe disease among residents and staff of long-term care facilities in Norway, November – June 2021

**DOI:** 10.1101/2021.08.08.21261357

**Authors:** Jostein Starrfelt, Anders S Danielsen, Oliver Kacelnik, Anita Wang Børseth, Elina Seppälä, Hinta Meijerink

## Abstract

COVID-19 has caused high morbidity and mortality in long-term care facilities (LTCFs) worldwide. We estimated vaccine effectiveness (VE) among residents and health care workers (HCWs) in LTCFs using Cox regressions. The VE against SARS-CoV-2 infection was 81.5 (95%CI: 75.3 – 86.1 82.7%) and 81.4% (95%CI: 74.5-86.4%) ≥ 7 days after 2^nd^ vaccine dose among residents and staff respectively. The VE against COVID-19 associated death was 93.1% among residents, no hospitalisations occurred among HCW ≥7 days after 2^nd^ dose.

## COVID-19 vaccination in long-term care facilities in Norway

High age is the main factor associated with severe outcomes for COVID-19. SARS-CoV-2 outbreaks in long-term care facilities (LTCFs) have caused a large burden of disease. To open society without excess mortality while preserving healthcare capacity, many countries, including Norway, prioritised vaccinating residents and staff of LTCFs [1-3]. This policy is dependent on a sufficient vaccine effectiveness (VE) in this population. Norway has the unique opportunity to follow both staff and residents for a complete picture of VE in this setting. The need for understanding VE is also vital for determining what kind of infection prevention and control measures are required. The balance between safety and quality of life is a fine line in LTCFs.Only a few studies have assessed the effect of COVID-19 vaccination among residents of LTCFs [1, 3-6]; showing that COVID-19 vaccines reduce both infections and severity of disease. Furthermore, a high VE in staff and residents is crucial for protecting individuals unable or unwilling to take the vaccine. In this cohort study, we estimate the effectiveness of COVID-19 vaccines in preventing PCR-confirmed SARS-CoV-2 infections in residents and staff of LTCFs, COVID-19 associated hospitalisation in staff and deaths in residents as a proxy for severe disease, to provide evidence for guidance and implementation of COVID-19 vaccination programmes.

### Determining vaccine effectiveness in long-term care facilities

We obtained data from BeredtC19, a preparedness registry containing individual-level data from various Norwegian registries (Supplementary table S1) [7]. We included data six weeks prior to vaccination start in Norway (27^th^ of December 2020), up to a PCR positive SARS-CoV-2 test, hospitalisation with COVID-19 as primary diagnosis or COVID-19 associated death, death from any cause, or end of follow-up (15th June 2021). We included all HCWs employed at LTCFs in the third week of January 2021 and residents registered with a long-term stay at a LTCF in 2020. We excluded those with prior SARS-CoV-2 infection, and individuals where the interval between first and second dose did not adhere to national recommendations [8, 9]. Underlying conditions were categorized as “high risk” or “medium risk” as stipulated by the national vaccination programme [10]. Vaccination status was defined as:

- Unvaccinated: unvaccinated, <14 days after 1^st^ vaccine dose
- Partly vaccinated_≥14 days after 1^st^ vaccine dose, <6 days after 2^nd^ vaccine dose
- Fully vaccinated: ≥7 days after 2^nd^ vaccine dose.

We included 31489 residents, of whom 85.4% (26905) received at least 1 dose during the follow-up period compared to 71.1% (63000) of the 88 549 HCWs. The median age was 87 years (IQR: 81-92 years) for residents and 39 (IQR: 27-53 years) for HCWs. Characteristics of the study population are shown in table 1. The incidence rates of COVID-19 outcomes were associated with vaccination status in both residents and HCWs (figure 1).

**Table 1.** Overview of characteristics of the study populations, namely residents and health care working in long-term care facilities (LTCF).

| Characteristics | Residents LTCF (31 489) |  | Health care workers LTCF (88 549) |  |
| --- | --- | --- | --- | --- |
|  | % | n | % | n |
| **Sex** |  |  |  |  |
| Female | 68.5 | 21 574 | 89.5 | 79 278 |
| Male | 31.5 | 9 915 | 10.5 | 9 271 |
| **Underlying conditions** |  |  |  |  |
| High | 45.1 | 14 192 | 12.2 | 10 822 |
| Medium | 8.5 | 2 677 | 1.0 | 847 |
| None | 46.4 | 14 620 | 86.8 | 76 880 |
| **Type of vaccine** |  |  |  |  |
| Comirnaty | 99.9 | 27 038 | 57.7 | 39 622 |
| Spikevax | 0.1 | 27 | 4.2 | 2 882 |
| Vaxzevria | <0,1 | <5 | 3.1 | 2 111 |
| Vaxzevria + mRNA* | <0,1 | <5 | 35.1 | 24 099 |

\* Vaxzevria was discontinued in Norway and those who received their first dose were offered a second dose of either Comirnaty or Spikevax

**Figure 1.**
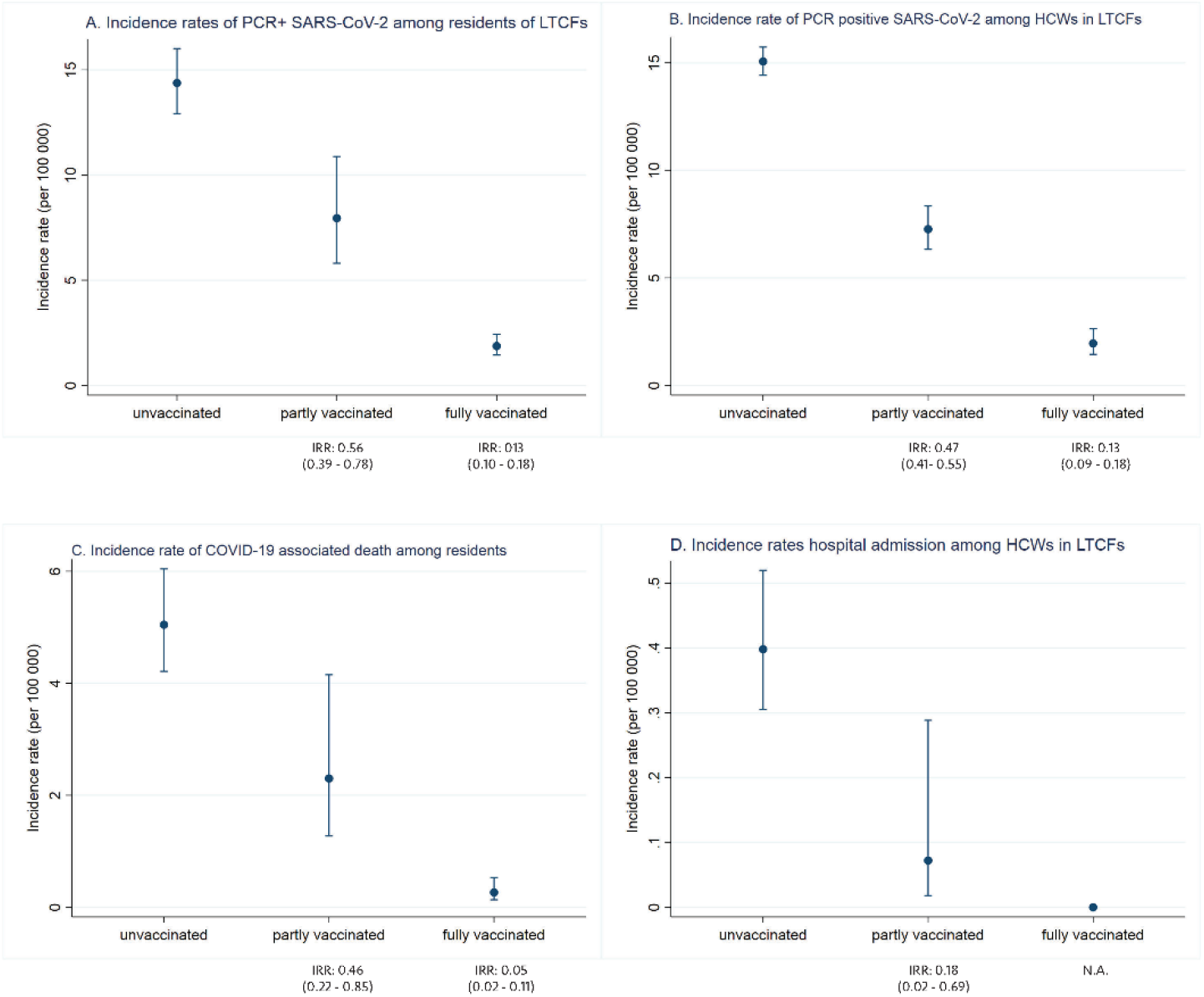
Incidence rates per 100,000 person-days with 95% confidence intervals by vaccination status for A) PCR positive SARS-CoV-2 infection and C) COVID-19 associated death among residents in long-term care facilities (LTCF), and B) PCR positive SARS-CoV-2 infection and D) hospital admissions with COVID-19 as main cause among health care workers in LTCF. Unvaccinated: Unvaccinated or up to 13 days after 1st dose, partly vaccinated: ≥14 days after 1st dose up to 6 days after 2nd dose, fully vaccinated: ≥7 days after 2nd dose. IRR: incidence rate ratio (compared to unvaccinated).

Using Cox proportional hazard models, we estimated the VE by modelling COVID-19 vaccination status as a time-varying covariate, adjusting for age, sex, and underlying conditions. The overall adjusted VE against PCR positive SARS-CoV-2 infection was 81.0% (95% CI: 76.5 – 84.6%) for fully vaccinated and 40.8% (95% CI: 31.8 – 48.5%) for partly vaccinated including residents and HCWs.

### Residents of long-term care facilities

The VE against PCR positive SARS-CoV-2 was 81.5% among fully vaccinated residents. Sex, age, and underlying conditions had limited effect on the estimates, probably due to the relatively uniform population characteristics (Table 2). Despite significantly lower incidence rates (figure 1), we could not show protection among partly vaccinated residents using cox proportional hazard models. This can be explained by the quick vaccination roll-out (90% had received their first dose by 20 January), and the limited time contributed as partly vaccinated since residents received their second dose three weeks after first. This effect may also be compounded by a high all-cause mortality.

**Table 2.** Estimated crude and adjusted COVID-19 vaccine effectiveness (VE) against laboratory confirmed SARS-CoV-2 infection and COVID-19 associated death among residents of long-term care facilities (LTCF).

| Vaccine status | Events | Person time<br>(days) | Rate<sup>§</sup> | Crude |  | Adjusted* |  |
| --- | --- | --- | --- | --- | --- | --- | --- |
|  |  |  | (per 100 000) | VE | 95% CI | VE | 95% CI |
| PCR positive SARS-CoV-2 |  |  |  |  |  |  |  |
| Unvaccinated | 332 | 2 338 969 | 14.2 | Ref |  | Ref |  |
| Partly vaccinated | 38 | 478 310 | 7.9 | -20.4 | -69.8 – 14.6 | -20.4 | -70.0 – 14.7 |
| Fully vaccinated | 57 | 3 015 338 | 1.9 | 81.7 | 75.6 – 86.3 | 81.5 | 75.3 – 86.1 |
| COVID-19 associated death |  |  |  |  |  |  |  |
| Unvaccinated | 118 | 2 338 969 | 5.0 | Ref |  | Ref |  |
| Partly vaccinated | 11 | 478 310 | 2.3 | 14.8 | -69.6 – 57.2 | 22.7 | -52.6 – 60.8 |
| Fully vaccinated | 8 | 3 015 338 | 0.3 | 90.4 | 76.6 – 95.5 | 93.1 | 85.1 – 96.8 |

Unvaccinated = Unvaccinated or up to 13 days after 1st dose, partly vaccinated = 14+ days after 1st dose up to 6 days after 2nd dose, fully vaccinated = 7+ days after 2nd dose. <sup>§</sup> per 100 000 person-days, \*adjusted for sex, age, and underlying conditions

In Norway, residents of LTCFs often receive health care at the facility and are not admitted to hospitals unless it is in the best interests of the resident. This population has the highest COVID-19 mortality, we therefore estimated the VE against COVID-19 associated death. 6 710 residents died during the study period, of whom 137 died with COVID-19. The VE against COVID-19 associated death was 93.1% among fully vaccinated individuals (table 2).

### Health care workers in long-term care facilities

Among HCWs, the VE against PCR positive SARS-CoV-2 was 45.0% among partly and 81.4% among fully vaccinated, adjusting for age, sex, underlying conditions, and calendar-time (Table 3). In Norway, COVID-19 associated mortality in the general population is low, therefore we used hospital admissions with COVID-19 as primary diagnosis as a measure of disease severity in HCWs. 56 individuals were hospitalised with COVID-19 as their primary diagnosis, of which two were among partly vaccinated and none fully vaccinated. The adjusted VE against COVID-19 hospitalisation was 81.7% for partly vaccinated HCWs in LTCFs (table 3).

**Table 3.** Estimated crude and adjusted COVID-19 vaccine effectiveness (VE) against laboratory confirmed SARS-CoV-2 and hospitalisation with COVID-19 as main cause among health care workers of long-term care facilities (LTCF).

| Vaccine status | Events | Person time<br>(days) | Rate<sup>§</sup> | Crude |  | Adjusted* |  |
| --- | --- | --- | --- | --- | --- | --- | --- |
|  |  |  | (per 100 000) | VE | 95% CI | VE | 95% CI |
| PCR positive SARS-CoV-2 |  |  |  |  |  |  |  |
| Unvaccinated | 2040 | 13 562 011 | 15.0 | Ref |  | Ref |  |
| Partly vaccinated | 197 | 2 771 252 | 7.1 | 47.3 | 38.7 – 55.7 | 45.0 | 35.3 – 53.2 |
| Fully vaccinated | 42 | 2 148 238 | 2.0 | 83.0 | 76.7 – 87.6 | 81.4 | 74.5 – 86.4 |
| Hospital admission with COVID-19 as main cause |  |  |  |  |  |  |  |
| Unvaccinated | 54 | 13 562 011 | 0.4 | Ref |  | Ref |  |
| Partly vaccinated | 2 | 2 771 252 | 0.1 | 80.1 | 14.2 – 95.4 | 81.7 | 21.0 – 95.8 |
| Fully vaccinated | 0 | 2 148 238 | NA | NA | NA | NA | NA |

Unvaccinated = Unvaccinated or up to 13 days after 1st dose, partly vaccinated = 14+ days after 1st dose up to 6 days after 2nd dose, fully vaccinated = 7+ days after 2nd dose.
<sup>§</sup> per 100 000 person-days
\*adjusted for sex, age, and underlying conditions. Set on calendar time

## Discussion and conclusion

We found a high VE against both SARS-CoV-2 infection and severe disease in residents and HCWs in LTCFs in Norway. Our estimates coincide with other studies looking at similar settings with high disease burden such as LTCFs [1-6, 11-13]. Importantly, we showed that one vaccine dose protected well against hospital admission among HCWs.

In Norway, residents of LTCFs were identified as very vulnerable and prioritized for vaccination. Vaccination roll-out in this group was quick and efficient [8]. Vaccination might be withheld for medical reasons, for example frailty or a history of severe allergic reactions to vaccines [14].

Persons with contraindications to vaccination are also those with the highest risk of severe outcomes after COVID-19, which could bias the VE estimate downwards. However, this bias seems to be limited, as can be seen from the lack of effect of age, sex, and underlying conditions on the VE.

We found that the VE against both SARS-CoV-2 infection and hospitalisation in HCWs was high also after one dose. The number of HCWs admitted to hospital in relation to COVID-19 is low but even lower post vaccination. Furthermore, the total protection for unvaccinated residents is significantly increased through staff vaccination. The effect of one dose also underlines the importance of ensuring temporary staff have received one dose prior to work in LTCFs.

Several methodological aspects should be considered when interpreting our data. Testing around cases in LTCFs has been extensive and continues for 10 days after the last case, reducing the risk of bias. As the time between the two vaccine doses was three weeks for residents, the follow-up time for partly vaccinated residents was limited and could explain why we didn’t demonstrate a significant effect of first vaccination, even on a background of reduced incidence per 100,000. A similar scenario was reported in Denmark [4].

The outcomes of this study are essential for the guidance of COVID-19 measures in LTCFs. Our results showcase the importance of quickly achieving full protection among the frail and elderly, and the effectiveness of one dose in healthcare workers. The pandemic has restricted the life of residents in LTCF. Through better knowledge, we can adjust COVID-19 guidelines for residents in LTCFs to fewer, targeted measures which might contribute to better quality of life. This knowledge is transferable to similar situations, like influenza, where vaccination of both residents and HCWs is of particular importance.

## Data Availability

Data will not be available

## Supplementary materials

**Table S1.**
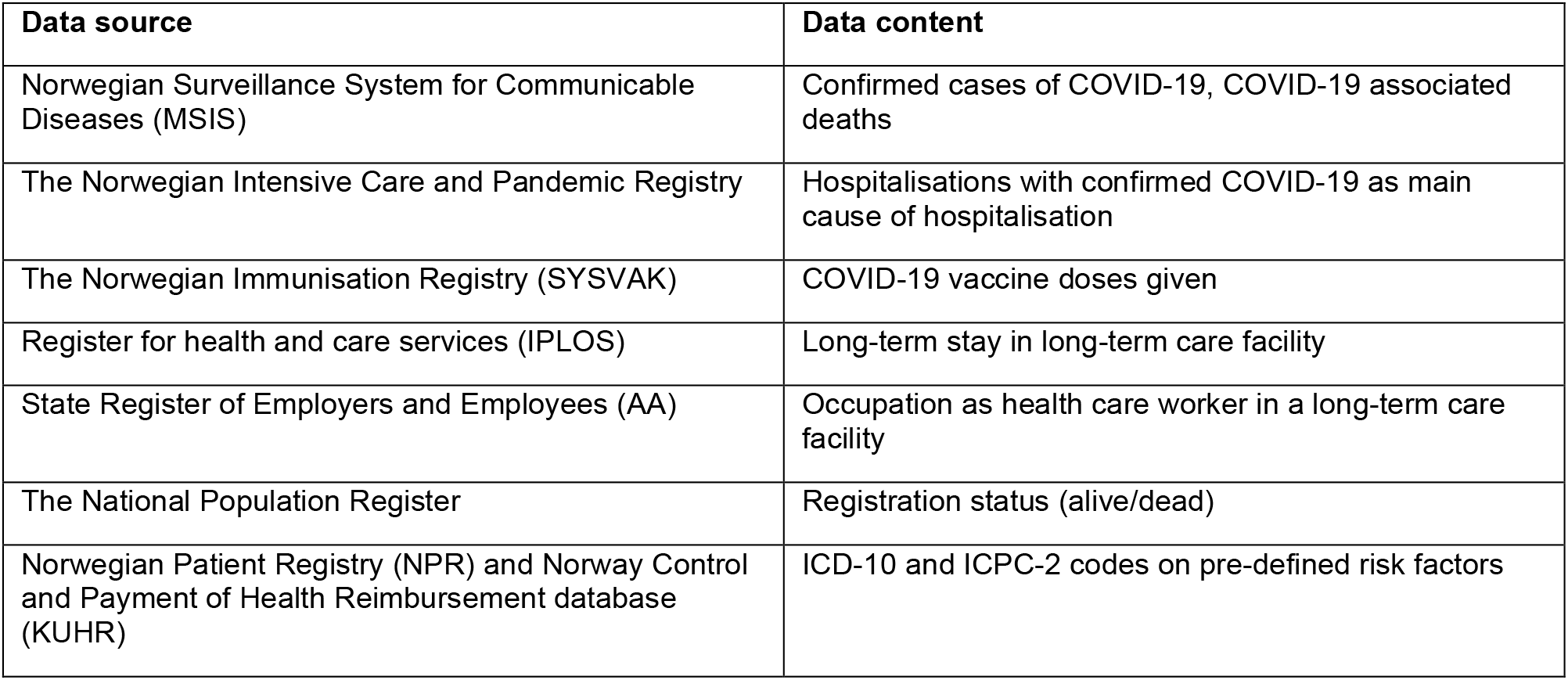
Data sources in the Norwegian preparedness registry BeredtC19 used in this study and individualized data retrieved from each source.

| Data source | Data content |
| --- | --- |
| Norwegian Surveillance System for Communicable Diseases (MSIS) | Confirmed cases of COVID-19, COVID-19 associated deaths |
| The Norwegian Intensive Care and Pandemic Registry | Hospitalisations with confirmed COVID-19 as main cause of hospitalisation |
| The Norwegian Immunisation Registry (SYSVAK) | COVID-19 vaccine doses given |
| Register for health and care services (IPLOS) | Long-term stay in long-term care facility |
| State Register of Employers and Employees (AA) | Occupation as health care worker in a long-term care facility |
| The National Population Register | Registration status (alive/dead) |
| Norwegian Patient Registry (NPR) and Norway Control and Payment of Health Reimbursement database (KUHR) | ICD-10 and ICPC-2 codes on pre-defined risk factors |

